# AImedReport: A Prototype Tool to Facilitate Research Reporting and Translation of AI Technologies in Healthcare

**DOI:** 10.1101/2024.01.16.24301358

**Authors:** Tracey A. Brereton, Momin M. Malik, Lauren M. Rost, Joshua W. Ohde, Lu Zheng, Kristelle A. Jose, Kevin J. Peterson, David Vidal, Mark A. Lifson, Joe Melnick, Bryce Flor, Jason D. Greenwood, Kyle Fisher, Shauna M. Overgaard

**Affiliations:** Center for Digital Health, Mayo Clinic, Rochester, Minnesota, United States of America; Department of Family Medicine, Mayo Clinic, Rochester, Minnesota, United States of America

## Abstract

AImedReport is a proof-of-concept team-based documentation strategy that consolidates available AI research reporting guidelines and centrally tracks and organizes any information provided as part of following various guidelines. It functions to assist teams by a) outlining phases of the AI lifecycle and clinical evaluation; b) iteratively developing a comprehensive documentation deliverable and historical archive; and c) addressing translation, implementation, and accountability gaps. By acting as a hub for determining what information to capture, it helps navigate team responsibilities, simplify compliance with evaluation and reporting measures, and fulfill requirements to support clinical trial documentation and publications. Here, we give an overview of this system and describe how it can be used to address documentation and collaboration challenges in AI translation.

## Introduction

The core of AI research in healthcare is carried out by AI data scientists, AI engineers, and clinicians; however, successfully evaluating and translating AI technologies into healthcare requires cross-collaboration beyond this group. Throughout ideation, development, and validation, successful translation requires engaging with many domains, including AI ethicists, quality management professionals, systems engineers, and more.^1-5^ We found through a scoping review that the prioritization of proactive evaluation of AI technologies, multidisciplinary collaboration, and adherence to investigation and validation protocols, transparency and traceability requirements, and guiding standards and frameworks are expected to help address present barriers to translation.^6^ However, as identified by Lu et al^7^ through a systematic review assessing clinical prediction model adherence to reporting guidelines that no consensus exists regarding model details that are essential to report, with some reporting items being commonly requested across reporting guidelines yet other reporting items being unique to specific reporting guidelines. Unless there is clear, consistent, and unified best practices and communication and collaboration across domains, there will be gaps in development, accountability, and implementation.^6-10^ Documentation is a crucial part of reporting and translation, but its coordinated maintenance throughout the AI lifecycle remains a challenge.^6,9-11^

We have established a proof-of-concept team-based documentation strategy for AI translation to simplify compliance with evaluation and research reporting standards through the development of AimedReport, a reporting guideline documentation repository. AimedReport organizes available reporting guidelines for different phases of the AI lifecycle, consolidating reporting items from different guidelines, assigning specific roles to team members, and guiding relevant information to capture when knowledge is generated (Appendix A).

**Figure 1.**
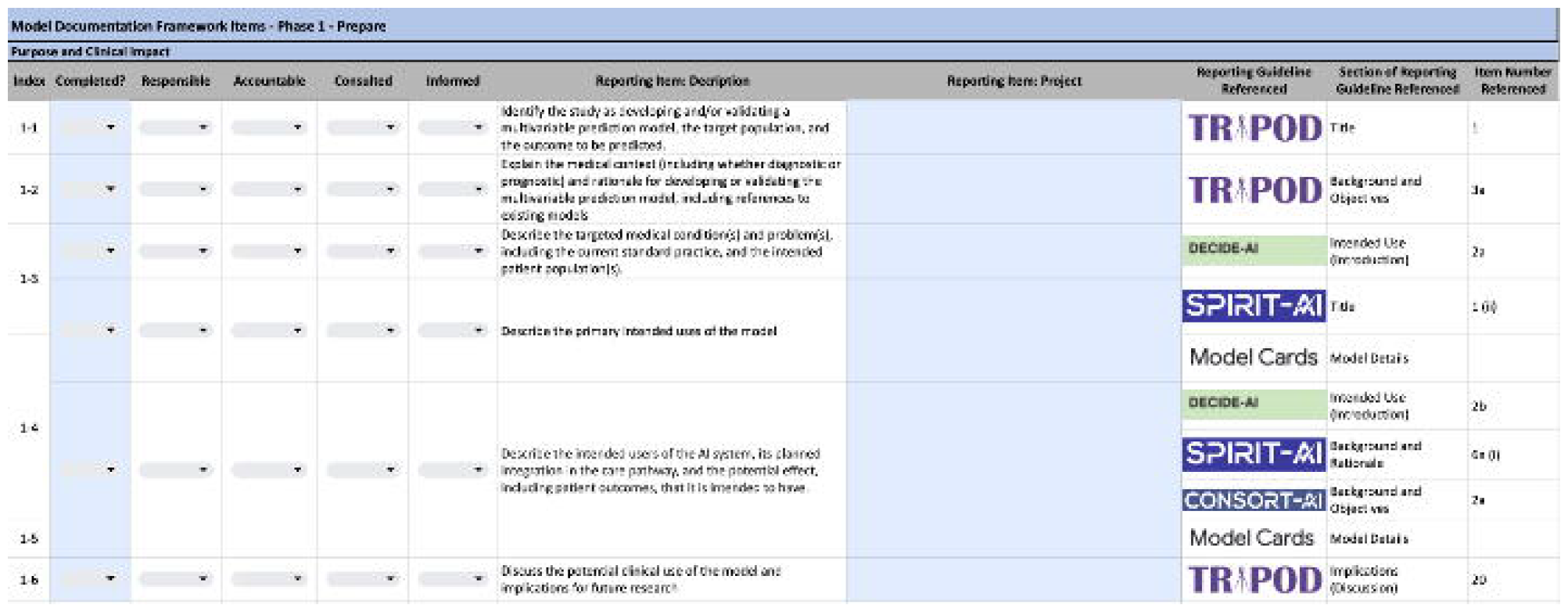
Prepare phase of AImedReport.

### Method of Development

We established a centralized documentation repository by first conducting a scoping review^6^ to investigate and understand the existing landscape of AI documentation and available resources (reporting guidelines, protocols, standards, frameworks, etc.). Within the scoping review, we found that documentation resources were fragmented throughout several reporting guidelines, prompting the consolidation and organization of such resources into AImedReport as a tool to structure available reporting guidelines in accordance with the AI lifecycle, reduce repetitive documentation burden, and promote knowledge continuity. Six research reporting guidelines make up the AImedReport, including: CONSORT-AI,^12^ DECIDE-AI,^13^ ML Test Score,^14^ Model Card,^15^ SPIRIT-AI,^16^ and TRIPOD^17^ (Table 1). The items that make up each reporting guideline are included in the AImedReport as “Reporting Items” and describe considerations for teams to document and maintain.

**Table 1.** Description of reporting guidelines included within the AImedReport.

| Reporting Guideline | Description |
| --- | --- |
| Consolidated Standards of Reporting Trials – Artificial Intelligence (CONSORT-AI) | Aims to promote transparency and completeness in reporting clinical trials for AI interventions, helping to understand, interpret and appraise the quality of clinical trial design and risk of bias in the reported outcomes; focuses on reporting the results of clinical trials |
| Developmental and Exploratory Clinical Investigation of DEcision-support systems driven by Artificial Intelligence (DECIDE-AI) | Aims to improve the reporting of studies describing the evaluation of AI-based decision support systems during their early, small-scale implementation in live clinical settings |
| ML Test Score | Aims to measure production readiness of a machine learning system is by offering a scoring system that focuses on assessing testing and monitoring needs |
| Model Card | Aims to encourage transparent model reporting, clarifying the intended use cases of models and detailing performance characteristics |
| Standard Protocol Items: Recommendations for Interventional Trials (SPIRIT-AI) | Aims to promote transparent prospective evaluation and completeness of clinical trial protocol reporting for AI interventions; focuses on publishing the clinical trial protocol before the trial is conducted |
| Transparent Reporting of a multivariable prediction model for Individual Prognosis or Diagnosis (TRIPOD) | Aims to improve the transparency and reporting of studies developing, validating, or improving a prediction model |

The AImedReport, conceptualized in 2022, was designed in concert with the AI Evaluation Framework by Overgaard et al.^1^ in 2022, which outlines clinical AI research and development stages. This alignment was established to support compliance with reporting standards and provide a reference for the entire AI development lifecycle, aiding in informing development phases, engaging stakeholders, and supporting interpretability, knowledge continuity, transparency, and trust. While based on the Overgaard et al.^1^ framework, AImedReport’s matured versatility allows it to potentially suit other frameworks such as van der Vegt’s^18^ SALIENT framework for broader AI implementation.

Each Reporting Item was mapped to one of the phases of the AI Evaluation Framework^1^ to streamline documentation when knowledge is generated: Prepare, Develop, Validate, Deploy, and Maintain. The “Prepare” phase focuses on metadata related to the owner, defining the model’s purpose and clinical impact, data preparation, and planning for model development. The “Develop” phase centers around model development and evaluation, usability related to inputs, assessing risk and bias, and protocol development for validation studies. The “Validate” phase catalogs information about the design and execution of how the model was validated, summative usability testing, generating user education, and planning for deployment. The “Deploy” phase focuses on clinical validation and generating training materials. Lastly, the “Maintain” phase plans for post-deployment surveillance and maintenance and quality monitoring and auditing. Reporting items were grouped into each of these five phases and then further classified into subgroups by identifying common themes (i.e., Prepare-Purpose and Clinical Impact; Develop-Model Development and Evaluation; Deploy-Clinical Validation). For each “Reporting Item”, the team or team members that need to be involved at each phase and in what ways (e.g., reporting, maintaining documentation, or utilization) were also defined, as shown in Table 2.

**Table 2.** AI lifecycle phases into which AImedReport “Reporting Items” were sorted, along with subphases and interdisciplinary alignment that came from further organization.

| Research & Discovery |  | Translation | Deployment |  |
| --- | --- | --- | --- | --- |
| Prepare | Develop | Validate | Deploy | Maintain |
| Purpose and Clinical Impact | Model Development and Evaluation | Validation Planning | Clinical Validation | Post-Deployment Surveillance and Maintenance |
| Data Preparation | Usability Formative | Usability Summative | User Training | Quality Monitoring and Audit |
|  | Model Bias Evaluation | User Education |  |  |
|  | Study Protocol Development | Deployment Planning |  |  |
| Project Manager | Project Manager | Project Manager | Project Manager | Clinical Expert |
| Clinical Expert | Clinical Expert | Clinical Expert | Clinical Expert | Quality Management |
| UX Researcher | UX Researcher | Ethicist | Data Scientist | MLOps |
| Ethicist | Ethicist | Data Engineer | MLOps | Ethicist |
| Data Engineer | Data Engineer | Data Scientist | System Engineer | Informaticist |
| Data Scientist | Data Scientist | System Engineer | Software Engineer | Software Engineer |
| Regulatory and Legal | Informaticist | UX Researcher | UX Researcher |  |
| Informaticist | System Engineer |  |  |  |
|  | Software Engineer |  |  |  |

## Discussion

The interactions among AI technologies, their users, and the implementation environments actively define the overall potential effectiveness of AI interventions within healthcare, especially because these tools are complex interventions designed as clinical decision support systems, not autonomous agents.^8,19,20^ A tailored, step-by-step approach may support the transition of AI technologies from being evaluated by statistical performance to clinical validity. To address this translational gap, AImedReport was developed to assist teams in several key areas, including a) outlining phases of the AI lifecycle and clinical evaluation, b) developing a comprehensive documentation deliverable and historical archive, and c) addressing translation, implementation, and accountability gaps. This is achieved by consolidating the existing landscape of research reporting guidelines into a repository. This repository acts as a centralized documentation hub and provides a standardized list of considerations and accountability assignments as the solution advances across the development lifecycle. AImedReport, accessible <u>here</u>, (Appendix A), is presently a prototype tool housed in a spreadsheet, but is planned to be made available as a Web resource and a software platform. This will likely further enhance the tool’s usability, reproducibility, and convenience by providing the ability to automate the documentation process, enhance task completion and generate deliverables in accordance with relevant reporting measures, and allow for communication and updates to model documents to be centrally available across teams.

Introducing such a platform allows for transparent communication of evaluation and reporting measures, but also embraces anticipated changes and modifications that come with development and maintenance. Each “Reporting Item” can be assigned to a team or team member to define who is responsible, accountable, consulted, and informed, who can then use the Reporting Item description as a reference to satisfy their role.^21^ For example, project managers, user experience researchers, and machine learning operations (MLOps) can contribute model overview, goals, and future state from their respective perspectives and reference one another’s vision. Similarly, data scientists, AI ethicists, informatics teams, and clinical practice committees may use documented demographic data of patient populations to assess items such as bias, differential model performance, appropriate clinical location, and potential clinical workflow location. During deployment and maintenance, the primary user and updater of the documentation will be an MLOps team, to ensure requirements set by previous groups are met, monitoring input and output metrics for drift, volume, and appropriate use. This can also facilitate interoperability between organizations, as the tool provides a standardized format, and documentation can be transferred across organizations and research governing bodies for consumption, auditing, and monitoring. AlmedReport also serves as a source of information describing completed evaluation and research reporting measures and can therefore fulfill reporting requirements to support clinical trial documentation and other publications. Additional descriptions of roles and responsibilities included within AImedReport can be found in Appendix B.

This paper focused on describing the theorization and development of AImedReport as a proof of concept to aid in evaluating, consolidating, and understanding available documentation resources to support AI reporting and facilitate communication across a multidisciplinary team. AlmedReport primarily concentrated on research reporting guidelines to address the immediate gaps identified within documentation practices. We note recent progress as the field rapidly advances towards enhancing implementation strategies within multidisciplinary teams. For example, in a study conducted by van der Vegt et al.^18^ titled “Implementation Frameworks for End-to-End Clinical AI: Derivation of the SALIENT Framework,” an extensive mapping exercise was conducted to synchronize various guidelines with an AI implementation framework. We suggest that AImedReport could further contribute to such implementation endeavors as a valuable resource. Planned future work will continue to converge with and align to various frameworks, like the SALIENT framework,^18^ ABCDS, ^22^ and organizations, including the Office of the National Coordinator,^23^ Food & Drug Administration,^24^ Coalition for Health AI,^25^ National Academy of Medicine,^5^ Health AI Partnership,^26^ National Institute of Standards and Technology,^27^ World Health Organization,^28^ and others.

We believe that AImedReport can be used in its current formative state for researchers and healthcare organizations to adhere to evaluation and research reporting standards, as well as to bridge some of the reporting and documentation requirements for products necessitating design controls under good manufacturing practices of the quality system regulation, such as those that may be Software as a Medical Device (SaMD).^28-30^ Future iterations of AlmedReport will better align translational science and regulatory science so that documentation can be used directly by teams pursuing regulated pathways, aligning with the information needed by regulatory review groups, accreditation commissions, and regulatory bodies (e.g., Food & Drug Administration).

### Next Steps and Conclusion

Our multidisciplinary team developed AImedReport as a strategic effort to address collaboration and documentation challenges in AI translation. AImedReport functions to assist teams by a) outlining phases of the AI lifecycle and clinical evaluation, b) iteratively developing a comprehensive documentation deliverable and historical archive, and c) addressing translation, implementation, and accountability gaps. By consolidating the existing landscape of research reporting guidelines into a repository, AImedReport acts as a centralized documentation hub that provides a standardized list of considerations and accountability assignments to guide information capture when knowledge is generated and simplify compliance with evaluation and reporting measures as AI technologies advances across the lifecycle. Completed measures documented within the AlmedReport may also serve as a source of information to fulfill reporting requirements to support clinical trial documentation and other publications. The integration of AImedReport into existing IT infrastructure and reporting platforms has undergone phased development, starting with the creation of a Model Documentation Framework presented at the AMIA 2022 Clinical Informatics Conference, refined through feedback from the Coalition for Health AI in 2022,^31^ and forming the foundation for collaborative efforts across various AI evaluation considerations. Mayo Clinic’s regulatory and systems engineering teams are adapting the AIMedReport framework to fit within regulatory infrastructure, aiming to scale multidisciplinary reporting for enterprise-wide AI applications.

This integration process involves continued interdisciplinary collaboration and evaluation to ensure scalability and applicability across Mayo Clinic departments and disciplines. Future work will include expanding AImedReport beyond a proof-of-concept phase and supporting various frameworks and organizations to enhance usability, including direct alignment of translational and regulatory sciences through FDA SaMD documentation.

## Data Availability

All data produced in the present work are contained in the manuscript.

https://docs.google.com/spreadsheets/d/1jenXP5miRxcteV6XRU71e-A7sJB46Ztz/edit#gid=603674031

## Abbreviations

AI: artificial intelligence
MLOps: machine learning operations
UX: user experience.

## Appendix

## Appendix A AImedReport

AImedReport is comprised of reporting items, outlined following product lifecycle phases to guide translation and promote transparent and explainable AI/ML-based MMS documentation. Access to the live document can be found here: AImedReport Link

## Appendix B Example AI Research Team Roles and Responsibilities

Appendix B: Example AI Research Team Roles and Responsibilities
| Role | Example Accountability |
| --- | --- |
| Project Manager | Manage the development and execution of a project, tracking progress and team communication to achieve project objectives. |
| Clinical Expert | Provides knowledge of clinical context throughout the AI lifecycle to inform design, usability, and adoption. |
| UX Researcher | Gathers user feedback and understands mental models of end users. Identifies users' needs and behaviors through direct observations, interviews, surveys, and usability evaluations to inform product design and development. |
| Ethicist | Identify types of possible harms, and specific potential subjects of harms; reason about underlying unjust historical and social processes that might eventually lead to differential model performance via measurement error, choice of sample, what data for which subjects are available or not. |
| Data Engineer | Manage data collection, connections, quality, cleaning, storage, processing and create data pipelines. |
| Data Scientist | Conduct data cleaning and quality checks, as well as employ appropriate statistical and machine learning approaches to uncover underlying meaningful relationships and patterns within datasets to allow for downstream actionable outcomes |
| MLOps | Actualize machine learning models into practice implementation and deployment through constructing necessary data connections and pipelines into production environments with appropriate model auditing, maintenance, monitoring, and feedback mechanisms in place. |
| System Engineer | Plan, define, test, and evaluate products for safety and efficacy against established best practices and procedures and external guidance or standards |
| Software Engineer | Build out data pipelines towards front-end users, accounting for resilient, scaled, and accurate cloud services through collaboration with data engineers and project developers to capture requirements and algorithms within application end products |
| Translational Informaticists | Bridges AI-model development and clinical implementation by aligning scientific and operational principles across teams |
| Regulatory and Legal | Manage and strategize compliance with regulatory policies and procedures, including those related to device classification and risk assessments. |
| Quality Management | Define and establish internal processes to fulfill quality mandates driven by institutions or external governance bodies. |

## References

1. Overgaard SM, Peterson KJ, Wi CI, et al. A Technical Performance Study and Proposed Systematic and Comprehensive Evaluation of an ML-based CDS Solution for Pediatric Asthma. AMIA Jt Summits Transl Sci Proc. 2022;2022:25–35.

2. Lysaght T, Lim HY, Xafis V, Ngiam KY. AI-Assisted Decision-making in Healthcare: The Application of an Ethics Framework for Big Data in Health and Research. Asian Bioeth Rev. Sep 2019;11(3):299–314. doi:10.1007/s41649-019-00096-0

3. IEEE. Addressing ethical dilemmas in AI: listening to engineers report. 2021. https://standards.ieee.org/initiatives/artificial-intelligence-systems/ethical-dilemmas-ai-report.html

4. Gilliland CT, White J, Gee B, et al. The Fundamental Characteristics of a Translational Scientist. ACS Pharmacol Transl Sci. Jun 14 2019;2(3):213–216. doi:10.1021/acsptsci.9b00022

5. Health Care Artificial Intelligence Code of Conduct. Accessed 12/1/2023. https://nam.edu/programs/value-science-driven-health-care/health-care-artificial-intelligence-code-of-conduct/

6. Brereton TA, Malik MM, Lifson M, Greenwood JD, Peterson KJ, Overgaard SM. The Role of Artificial Intelligence Model Documentation in Translational Science: Scoping Review. Interact J Med Res. Jul 14 2023;12:e45903. doi:10.2196/45903

7. Lu JH, Callahan A, Patel BS, et al. Assessment of Adherence to Reporting Guidelines by Commonly Used Clinical Prediction Models From a Single Vendor: A Systematic Review. JAMA Netw Open. Aug 1 2022;5(8):e2227779. doi:10.1001/jamanetworkopen.2022.27779

8. Seneviratne MG, Shah NH, Chu L. Bridging the implementation gap of machine learning in healthcare. Bmj Innov. Apr 2020;6(2):45–47. doi:10.1136/bmjinnov-2019-000359

9. DeCamp M, Lindvall C. Latent bias and the implementation of artificial intelligence in medicine. J Am Med Inform Assoc. Dec 9 2020;27(12):2020–2023. doi:10.1093/jamia/ocaa094

10. Amann J, Blasimme A, Vayena E, Frey D, Madai VI. Explainability for artificial intelligence in healthcare: a multidisciplinary perspective. BMC Med Inform Decis Mak. Nov 30 2020;20(1):310. doi:10.1186/s12911-020-01332-6

11. Andrews E. “Flying in the dark”: Hospital AI tools aren’t well documented. 2021. Accessed 2/1/2022. https://hai.stanford.edu/news/flying-dark-hospital-ai-tools-arent-well-documented

12. Liu X, Cruz Rivera S, Moher D, et al. Reporting guidelines for clinical trial reports for interventions involving artificial intelligence: the CONSORT-AI extension. Lancet Digit Health. Oct 2020;2(10):e537–e548. doi:10.1016/S2589-7500(20)30218-1

13. Vasey B, Nagendran M, Campbell B, et al. Reporting guideline for the early-stage clinical evaluation of decision support systems driven by artificial intelligence: DECIDE-AI. Nat Med. May 2022;28(5):924–933. doi:10.1038/s41591-022-01772-9

14. Breck E, Cai S, Nielsen E, Salib M, Sculley D. The ML test score: A rubric for ML production readiness and technical debt reduction. IEEE International Conference on Big Data (Big Data). 2017:1123–1132. doi:10.1109/BigData.2017.8258038.

15. Mitchell M, Wu S, Zaldivar A, et al. Model Cards for Model Reporting. Proceedings of the conference on fairness, accountability, and transparency. 2019:220–229. doi:10.1145/3287560.3287596

16. Cruz Rivera S, Liu X, Chan AW, et al. Guidelines for clinical trial protocols for interventions involving artificial intelligence: the SPIRIT-AI extension. Lancet Digit Health. Oct 2020;2(10):e549–e560. doi:10.1016/S2589-7500(20)30219-3

17. Collins GS, Dhiman P, Andaur Navarro CL, et al. Protocol for development of a reporting guideline (TRIPOD-AI) and risk of bias tool (PROBAST-AI) for diagnostic and prognostic prediction model studies based on artificial intelligence. BMJ Open. Jul 9 2021;11(7):e048008. doi:10.1136/bmjopen-2020-048008

18. van der Vegt AH, Scott IA, Dermawan K, Schnetler RJ, Kalke VR, Lane PJ. Implementation frameworks for end-to-end clinical AI: derivation of the SALIENT framework. J Am Med Inform Assoc. Aug 18 2023;30(9):1503–1515. doi:10.1093/jamia/ocad088

19. Grote T, Berens P. How competitors become collaborators-Bridging the gap(s) between machine learning algorithms and clinicians. Bioethics. Oct 2 2021;doi:10.1111/bioe.12957

20. Li RC, Asch SM, Shah NIH. Developing a delivery science for artificial intelligence in healthcare. Npj Digital Medicine. Aug 21 2020;3(1)doi:ARTN 107 10.1038/s41746-020-00318-y

21. Brower HH, Nicklas BJ, Nader MA, Trost LM, Miller DP. Creating effective academic research teams: Two tools borrowed from business practice. J Clin Transl Sci. Nov 5 2020;5(1):e74. doi:10.1017/cts.2020.553

22. Bedoya AD, Economou-Zavlanos NJ, Goldstein BA, et al. A framework for the oversight and local deployment of safe and high-quality prediction models. J Am Med Inform Assoc. Aug 16 2022;29(9):1631–1636. doi:10.1093/jamia/ocac078

23. healthIT.gov. Clinical Decision Support. 2018;

24. Accessed 12/1/2023, https://www.fda.gov/medical-devices

25. AI CfH. Accessed 12/1/2023, https://coalitionforhealthai.org/

26. Partnership HA. Accessed 12/1/2023, https://healthaipartnership.org/

27. AI RISK MANAGEMENT FRAMEWORK. Accessed 11/1/2023, https://www.nist.gov/itl/ai-risk-management-framework/ai-rmf-development

28. FG-AI4H deliverables overview. Accessed 5/1/2023, https://www.itu.int/en/ITU-T/focusgroups/ai4h/Pages/deliverables.aspx

29. CFR - Code of Federal Regulations Title 21. Accessed 11/1/2023, https://www.accessdata.fda.gov/scripts/cdrh/cfdocs/cfcfr/CFRSearch.cfm?fr=56.115

30. ISO/IEC JTC 1/SC 42. Accessed 12/1/2023, https://www.iso.org/committee/6794475.html

31. AI CfH. PROVIDING GUIDELINES FOR THE RESPONSIBLE USE OF AI IN HEALTHCARE. 2022. https://www.coalitionforhealthai.org/papers/Virtual%20Working%20Group%20Session%202%20-%20Testing,%20Usability%20and%20Safety.pdf

